# Food insecurity and cholera risk: An Exploratory Analysis of Potential Mediators

**DOI:** 10.1101/2022.06.13.22276311

**Authors:** Ahmed D. Elnaiem, Molly F. Franke, Aaron Richterman, Yodeline Guillaume, Kenia Vissieres, Gertrude Cene, Ralph Ternier, Louise C. Ivers

**Author notes:** ADE, MFF, LCI, AR, & YG. These authors contributed equally to the following: (1) conception and design of the study, acquisition of data, or analysis and interpretation of results; (2) writing, reviewing and the manuscript critically for intellectual content; and (3) final approval of the version submitted. KV, GC, & RT. These authors contributed equally to the following: (1) conception and design of the study, acquisition of data, or analysis and interpretation of results; (2) final approval of the version submitted.

## Abstract

**Background:** Food insecurity has been independently associated with cholera infection and there is an inverse relationship between national food security and annual cholera incidence. However, factors that mediate increased cholera risk among food insecure households remain largely unexplored.

**Methods:** In a cross-sectional survey of rural households in Haiti, we explored the role of food behaviors (i.e., dietary choices and food-handling practices) as mediators of cholera risk among food-insecure families. We generated multivariable regression models to test hypothesized associations between severity of food insecurity (measured by the Household Hunger Scale), hygiene and food behaviors, and history of severe, medically-attended cholera.

**Results:** Compared with little to no household hunger, moderate hunger (Adjusted Odds Ratio [AOR] 1.62, 95% Confidence Interval (CI) 1.12—2.36; p=0.011) and severe hunger (AOR 2.32, 95% CI 1.27—4.22; p=0.006) were positively associated with history of severe, medically-attended cholera. Household hunger was positively associated with three behaviors: antacid use, consumption of leftover non-reheated food, and eating food and beverages prepared outside of the home (i.e., at a restaurant or from a vendor). Consumption of outside food items and antacid use were positively associated with a history of cholera.

**Conclusion:** Our findings suggest that food behaviors may mediate the association between food insecurity and cholera and contribute to an understanding of how interventions could be designed to target food insecurity as part of cholera prevention and control.

**Author summary:** Food insecurity has been found to be a risk factor for cholera at the household and national level;[1–3] however the mechanism through which food insecurity may increase the risk of cholera remains unknown.

In a large cross-sectional survey of 1072 households in rural Haiti, we observed a robust independent association between food insecurity—defined as a persistent lack of access to food in adequate quantity or quality and measured by the Household Hunger Scale—and severe, medically-attended cholera. This relationship appears to be linear, conferring a dose-dependent risk of cholera by severity of food insecurity. We found household food insecurity to be associated with three high-risk behaviors: antacid use, consumption of leftover non-reheated food, and eating food and beverages prepared outside of the home. Two high-risk behaviors—including antacid use and consumption food and beverages prepared outside of the home (i.e., at a restaurant or from a vendor)—were independent risk factors for cholera.

High-risk food handling practices may be one causal pathway whereby food insecurity increases risk of cholera infection. Future longitudinal and qualitative studies should investigate whether interventions targeting food insecurity could reduce cholera risk among populations who face a high burden of both conditions.

## Introduction

Cholera and food insecurity are afflictions of inequity that sicken and kill impoverished and vulnerable populations. In 2018, cholera resulted in an estimated 1.3 to 4.0 million cases and 21,000 to 143,000 deaths worldwide.[4] Meanwhile, in the same year, an estimated 2 billion people faced moderate or severe food insecurity.[5] Close links between determinants of access to food, clean water, and basic sanitation give rise to significant overlaps in the distribution of food insecurity and cholera in impoverished communities.

Food insecurity is a multidimensional phenomenon defined by an *uncertain* or *limited availability* of nutritionally adequate or safe food.[6] Over the past two decades, a large and growing body of literature has identified direct relationships between food insecurity and adverse outcomes in metabolic disease,[7–9] infectious diseases (HIV, tuberculosis),[10] cardiovascular disease,[11–13] liver disease,[10,14] sleep disorders,[15] and mental health conditions (depression, anxiety, and psychological distress),[16] among others.[17]

Because of the well-documented relationship between food insecurity and poor health outcomes in other contexts, and the frequent co-occurrence of cholera and food insecurity in impoverished settings, we have recently begun to evaluate the potential role of food security in cholera risk. We previously found food insecurity to be an independent risk factor for both history of and death from cholera.[2,3] An analysis of 30 countries demonstrated a strong inverse relationship between national food security and the annual incidence of cholera.[1] Food insecurity may affect cholera outcomes through several hypothesized pathways: impaired immune and gut barrier function from malnutrition; suboptimal food hygiene practices resulting from altered risk-calculation because of increased attention to immediate food needs; and poor mental health undermining resilience to the economic losses and emotional stress imparted by cholera infection.[6,11,18,19] Yet, no studies to date have sought to elucidate which of these pathways in fact operates to increase cholera risk.

To address this knowledge gap, we assessed the potential role of household food behaviors (i.e., dietary choices and food-handling practices) as mediators of the relationship between food insecurity and cholera using a cross-sectional survey of rural Haitian households. We hypothesized that food insecurity would be positively associated with household behaviors that predispose to cholera, and these behaviors would be positively associated with cholera.

## Methods

### Study design, population, and measures

We analyzed data from a cross-sectional household survey administered to a random sample of 1072 households in Mirebalais, Haiti in 2019. The survey was conducted as part of a program to prevent and control cholera. We defined a household as an individual or group of related or unrelated individuals sleeping at least half the week under the same roof and sharing resources (i.e., food from the same pot or cost of living expenses). Respondents were consenting adult heads of the household (18 years or older) and, if unavailable, other adult household members capable of answering the survey questions about the household.

Household-level interviews captured socioeconomic characteristics, food insecurity, risk factors for cholera exposure, cholera vaccination status of household members, knowledge and practices related to cholera and water, sanitation, and hygiene (WASH). Survey respondents were asked about their diet, medication use, and food-handling practices over the week and month prior to interview. Study workers observed household water storage and treatment practices, handwashing spaces, toilet facilities, and household cleanliness (i.e., presence of trash in the yard or garden areas). During the observation phase of the interview, study workers also tested household drinking water samples for free-residual chlorine (a component of water quality) using a DPD (diethyl paraphenylenediamine) indicator test. We defined effective water treatment as free-residual chlorine level between 0.2 – 2 mg/L, per USAID guidelines.[20] Household food insecurity was measured using the Household Hunger Scale (HHS), a cross-culturally validated indicator consisting of three items drawn from the Household Food Insecurity Access Scale (HFIAS). Households were classified into three categories based on established thresholds: little to no hunger, moderate hunger, and severe hunger in the household.[21] History of severe, medically attended cholera was obtained by self-report, through two questions: “Has anyone in your household (besides you) ever spent the night in a cholera treatment unit?” and “Have you had diarrhea requiring you to stay overnight in a cholera treatment center or hospital since 2010*?*” We estimated household poverty status using the Simple Poverty Scorecard—a validated poverty assessment tool specific to Haiti based on 11 indicators.[22] The scorecard generates a numeric poverty score (continuous variable) and estimates the likelihood of consumption below a poverty line (about $1/day)—a lower score indicates higher relative poverty. All instruments were designed, translated, and back-translated by bilingual study team members and extensively field-tested in the study population in central Haiti.[3,23]

### Statistical analysis and causal framework

Our statistical analysis focused on the association between food insecurity (exposure) and household history of cholera (primary outcome) and an exploratory analysis of high-risk household behaviors as potential mediators of the association.

We first constructed a directed acyclic graph, based on published risk factors for cholera and biological plausibility, to guide our exploratory analysis and illustrate our hypothesized causal framework (Figure 1). We used logistic regression models to assess the relationship between household food insecurity and history of cholera. We generated an initial univariate model to calculate the crude unadjusted odds ratio (OR) with a 95% confidence interval (CI) between household hunger—the Household Hunger Scale modeled as moderate and severe food insecurity versus little to none[21]—and history of severe, medically-attended cholera in the household. We then performed multivariable analyses to calculate an adjusted odds ratio (AOR) and 95% CI.

**Fig. 1.**
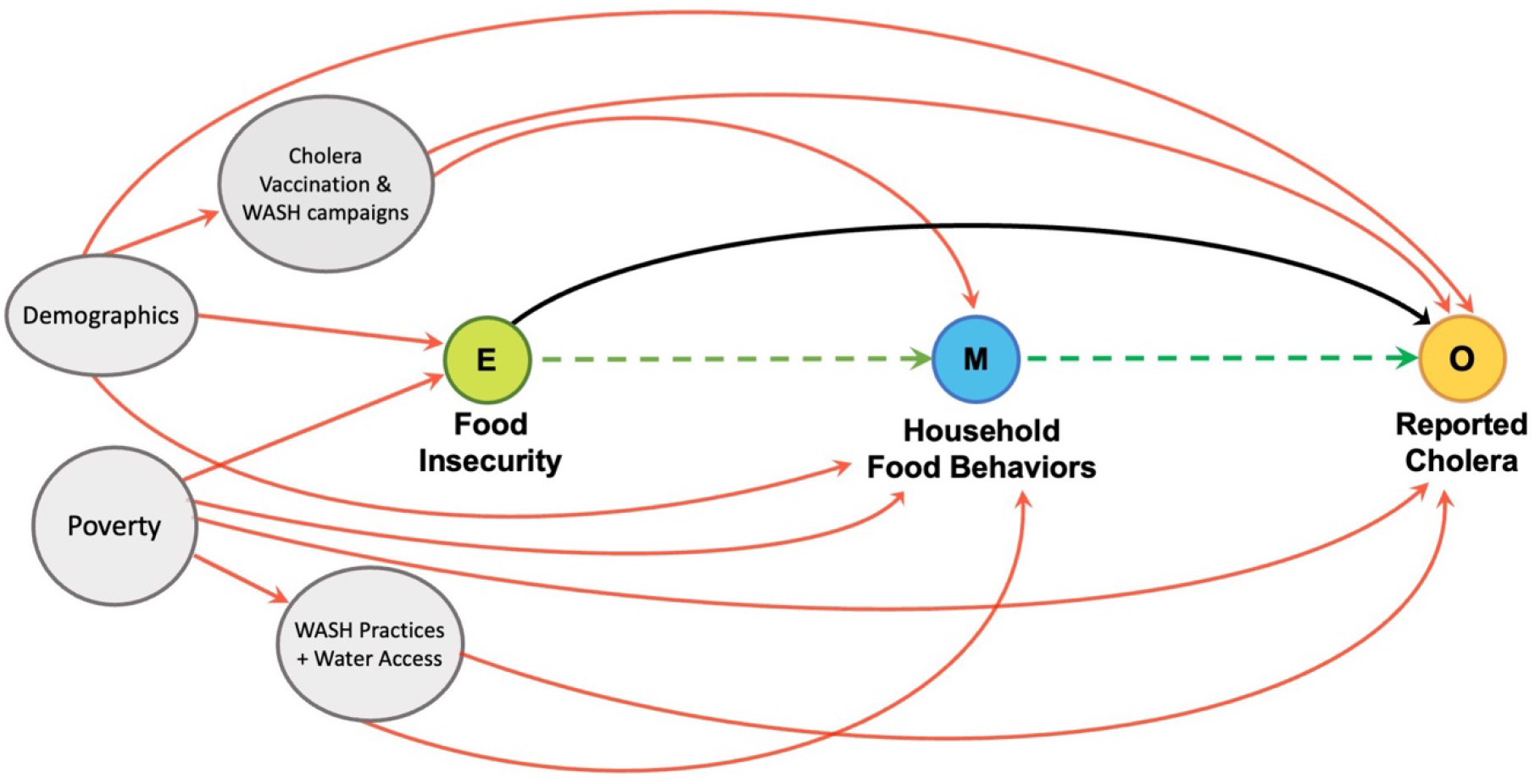
Directed acyclic graph illustrating hypothesized relationship pathways. Exposure is shown in green, mediating food behaviors in light blue, confounders in grey, and outcome in yellow. The indirect effect of food insecurity occurring through mediating household food behaviors is shown in green (dashed line). The direct of food insecurity on cholera is shown in black. Confounders are shown in grey with their effect in red.

Confounders, selected based on our causal framework, prior publications of risk factors for cholera, and observed associations in our data included: poverty score, single marital status (head of household), cholera vaccination status, improved household water source, money spent on water (0, 0-15, 15-30, and >30 Haitian gourdes), distance to a water source (<15 min, 15-30 min, 30-45min, 45 min – 1 hour, and >1 hour) and latrine use. [3,24–26] Of note, we opted to treat prior cholera vaccination and knowledge of handwashing as confounders of cholera given evidence from a previous longitudinal study which demonstrated cholera vaccination campaigns in a neighboring department were associated with increased handwashing knowledge and practice in part as a consequence of the vaccination campaign’s comprehensive messaging.[26] In a secondary analysis, we examined the household hunger scale as a linear variable, rather than categorical, and performed a log-likelihood ratio test to assess whether a linear, dose-dependent relationship better characterized the observed association between food insecurity and severe, medically-attended cholera.

To interrogate mediating effects through our hypothesized food behavior pathway, we adopted a two-step approach using logistic regression models to identify whether food insecurity was associated with specific high-risk behaviors in a household and whether these behaviors were associated with cholera. First, we used multivariate regression models to estimate the association between food insecurity and specific behaviors. We considered five variables: consumption of leftover unheated food, raw fruits/vegetables, raw or undercooked fish, foods prepared outside the home, and antacid use over the prior week. We ran separate multivariable regression models, with each of the five food behavior variables as the dependent variable, household food insecurity as the independent variable, and the following confounders based on our causal framework and observed associations in the data: poverty score, single marital status, prior cholera vaccination, and knowledge of handwashing.

We then used multivariate regression models to assess the association between specific food behavior variables and history of household cholera. We adjusted for the same cholera risk factors included in the overall model estimating the association between food insecurity and cholera. To evaluate for multicollinearity in multivariate models, we calculated a variance inflation factor among covariates for each model—a factor greater than 2.50 was deemed indicative of multicollinearity.

We conducted a complete case analysis. Missing data was present for some variables collected during study worker observations of household WASH infrastructure. To assess for potential bias arising from missing observations for variables that were strongly associated with both cholera and food insecurity, we conducted a sensitivity analysis employing missing indicator variables to test whether exclusion of missing observations in our complete case analysis introduced bias into the models.

All quantitative analyses were performed using R Studio version 1.2.1335 for Mac with R version 3.6.1 for Mac. Institutional Review Board approval was received from Partners Healthcare (Protocol #2018P001220) & Zanmi Lasante, Haiti (Protocol #128).

## Results

One thousand one hundred twenty households were selected to participate in the study, of which 9 declined to participate, 28 were not contactable, and 20 did not reside permanently in the study’s catchment area. A total of 1072 households, with a median size of 5 individuals (IQR 3-6), consented to participate in the survey (Table 1). Among all respondents, 52% were female, 60% worked primarily in agriculture, 42% never attended school, and 27% were single-parent heads of household. Approximately one-third of households (n = 343) reported at least one instance of severe, medically-attended cholera. Most households (64.4%) experienced either moderate or severe levels of household hunger.

**Table 1.** Household characteristics, dietary, food, and hygiene practices and knowledge of cholera in a cross-sectional household survey of Mirebalais, Haiti 2019.

|  |  | Little to no Hunger **<br>N = 382 |  | Moderate Hunger **<br>N = 598 |  | Severe Hunger **<br>N=92 |  |
| --- | --- | --- | --- | --- | --- | --- | --- |
|  |  | n | % | n | % | n | % |
| **Household Characteristics** |  |  |  |  |  |  |  |
| Household size [median (25<sup>th</sup>-75<sup>th</sup> percentile)] |  | 4 (3 - 6) |  | 5 (3 - 6) |  | 5 (4 - 7) |  |
| Number of children < 5 years old |  | 108 | 28% | 213 | 36% | 42 | 46% |
| Single marital status (head of household) |  | 120 | 31% | 203 | 34% | 34 | 37% |
| Poverty score [median (standard deviation)] |  | 48 (15.9) |  | 40 (15.1) |  | 31 (14) |  |
| Likelihood of consumption below national poverty line [22] |  | 44.8% |  | 62.7% |  | 83.6% |  |
| Household history of severe, medically-attended cholera |  | 85 | 22% | 211 | 35% | 47 | 51% |
| **Dietary and Other Intake Practices\*** |  |  |  |  |  |  |  |
| Antacid use |  | 174 | 46% | 258 | 43% | 61 | 66% |
| Family was unable to reheat leftover food before eating |  | 78 | 20% | 121 | 20% | 43 | 47% |
| Consumed food/beverage prepared outside of the home |  | 121 | 32% | 280 | 47% | 60 | 65% |
| Source of procured item(s) | Family/friend's house | 48 | 40% | 162 | 58% | 46 | 77% |
|  | Restaurant | 35 | 29% | 68 | 24% | 6 | 10% |
|  | Marketplace | 37 | 31% | 99 | 35% | 32 | 53% |
| Dietary composition and medication use | Raw fruits and vegetables | 210 | 55% | 271 | 45% | 40 | 43% |
|  | Raw or undercooked fish | 11 | 3% | 17 | 3% | 5 | 5% |
|  | Almost daily fish or seafood consumption | 27 | 38% | 12 | 55% | 3 | 73% |
|  | Leftover unheated rice | 71 | 19% | 124 | 21% | 48 | 52% |
|  | Beverage with ice obtained outside of household | 288 | 75% | 407 | 68% | 76 | 83% |
| **Water, Sanitation and Hygiene Knowledge and Reported Practices** |  |  |  |  |  |  |  |
| Utilization of an improved household water source<sup>†</sup> |  | 190 | 50% | 230 | 38% | 30 | 33% |
| Water from usual source was unavailable at least 1 whole day in the past month |  | 153 | 40% | 326 | 55% | 44 | 48% |
| Always treats household water |  | 272 | 71% | 247 | 41% | 30 | 33% |
| Reason for not always treating water | Too expensive | 4 | 1% | 54 | 15% | 0 | 0% |
|  | We don't have what we need to treat | 54 | 49% | 208 | 59% | 46 | 74% |
|  | Hard to get what we need to treat | 27 | 25% | 114 | 32% | 35 | 56% |
| Time to fetch water and return by foot | <15 min | 204 | 53% | 234 | 39% | 40 | 43% |
|  | 15-29 min | 69 | 18% | 128 | 21% | 24 | 26% |
|  | 30-59 min | 58 | 15% | 160 | 27% | 16 | 17% |
|  | 1 hour + | 51 | 13% | 76 | 13% | 12 | 13% |
| Money spent daily on water # | None | 192 | 50% | 440 | 74% | 73 | 79% |
|  | 0 - 30 gourdes | 119 | 31% | 84 | 14% | 9 | 10% |
|  | > 30 gourdes | 71 | 19% | 74 | 12% | 10 | 11% |
| Washes hands with soap and water ≥4 times daily |  | 261 | 68% | 280 | 47% | 68 | 74% |
| Soap always available in the home |  | 241 | 63% | 206 | 34% | 29 | 32% |
| Offers this example of when hand washing is appropriate | After the toilet | 262 | 69% | 476 | 80% | 77 | 84% |
|  | Before eating | 332 | 87% | 517 | 86% | 78 | 85% |
|  | Before preparing food | 166 | 43% | 170 | 28% | 22 | 24% |
|  | After touching things like money telephones, etc. | 147 | 38% | 119 | 20% | 30 | 33% |
|  | After handling a person with vomiting or diarrhea | 71 | 19% | 82 | 14% | 9 | 10% |

\* Primary survey respondents (head of household) were asked about consumption over the prior week.
\*\* Household Hunger Scale scored as: little to no hunger in the household (score 0-1), moderate hunger in the household (score 2-3), or severe hunger in the household (score 4-6).
† Improved water sources were defined by Joint Monitoring Programme for Water Supply and Sanitation (JMP) standards and included: piped household water, protected wells or springs, bottled water, and collected rainwater. [32]

The distribution of socioeconomic characteristics, food behaviors, and WASH knowledge and reported practices varied significantly by the severity of household hunger (Table 1). The week before interview, moderately and severely hungry households were more likely to have consumed food or beverages prepared outside and been unable to reheat their food. Households with severe hunger reported more frequent use of antacids (66% vs. 43% in moderate, and 46% in little to no hunger; p<0.0001) and were least likely to report always treating their water (33%, p<0.0001). Households with moderate and severe hunger were less likely to spend money on water (p<0.0001), collect from an improved water source (p = 0.0003), or travel less than 15 minutes to fetch water and return (p<0.0001). These households were also more likely to store water in a wide-mouthed container in which a hand could fit (66% for moderate; 61% for severe) compared to those with little to no hunger (48%) (Table 2).

**Table 2.** Study worker observations of household water storage and treatment practices, handwashing spaces, toilet facilities, and environmental hygiene in a cross-sectional household survey of Mirebalais, Haiti 2019.

|  |  | Little to no Hunger<br>N = 382 |  | Moderate Hunger<br>N = 598 |  | Severe Hunger<br>N = 92 |  | Missing* |
| --- | --- | --- | --- | --- | --- | --- | --- | --- |
| Household Observations |  | n | % | n | % | n | % | n |
| Free residual chlorine measured in drinking water, mean (95% CI) mg/L |  | 0.3 (0.2 - 0.4) |  | 0.6 (0.5 - 0.7) |  | 0.8 (0.6 - 1.1) |  | 284 |
|  | Within effective range (0.2 - 2 mg/L) | 50 | 19% | 128 | 29% | 31 | 40% |  |
|  | Below effective range (<0.2 mg/L) | 212 | 79% | 288 | 65% | 38 | 49% |  |
| Water treatment products visible on the household premises |  | 259 | 68% | 469 | 83% | 69 | 75% |  |
| Use of covered water storage vessel |  | 204 | 85% | 380 | 81% | 61 | 79% | 282 |
| Opening of water storage vessel | Narrow-mouthed (hand cannot fit in) | 125 | 52% | 163 | 35% | 30 | 39% |  |
|  | Wide-mouthed (hand can fit in) | 116 | 48% | 309 | 66% | 47 | 61% |  |
| Method of drawing water from | With dedicated ladle or utensil | 193 | 80% | 349 | 74% | 64 | 83% |  |
| container | With non-dedicated utensil | 19 | 8% | 84 | 18% | 24 | 31% |  |
| No dedicated household handwashing space |  | 353 | 92% | 573 | 96% | 85 | 92% | 12 |
| Loose rubbish visible in yard/garden area |  | 90 | 24% | 297 | 50% | 50 | 54% |  |
| Household toilet system | Flushing toilet | 48 | 17% | 13 | 3% | 3 | 4% | 244 |
|  | Improved pit latrine | 106 | 38% | 120 | 25% | 9 | 12% |  |
|  | Unimproved pit latrine | 112 | 40% | 268 | 56% | 53 | 73% |  |
|  | Open area or garden | 10 | 4% | 60 | 13% | 7 | 10% |  |
|  | Compositing toilet or bucket | 4 | 1% | 14 | 3% | 1 | 1% |  |
| Where a toilet system was present, fecal matter seen on exposed surface |  | 49 | 18% | 153 | 32% | 21 | 29% |  |
| Number of household members sharing toilet, mean (95% CI) |  | 1.8 (1.7 - 2.1) |  | 2.0 (1.8 - 2.1) |  | 2.3 (1.5 -3.1) |  | 321 |
| Flies visible in latrine |  | 31 | 12% | 102 | 28% | 15 | 25% | 397 |

\* Missing observations reflect instances where respondents declined observation of household facilities or study workers were unable to complete the observation portion of the interview due to other extenuating circumstances.

### Association of Food Insecurity and Cholera

Univariate analysis demonstrated a significant association between moderate (OR 1.91, 95% CI 1.42 - 2 .56; p<0.001) and severe (OR 3.65, 95% CI 2.27 - 5.88; p<0.001) household hunger and history of cholera relative to little to no household hunger (Table 3). Multivariable analysis including hypothesized confounders yielded a similar, though attenuated association for moderate (AOR 1.62, 95% CI 1.12-2.36; p=0.01) and severe (AOR 2.32, 95% CI 1.27-4.22; p=0.006) hunger in the household.

**Table 3.**
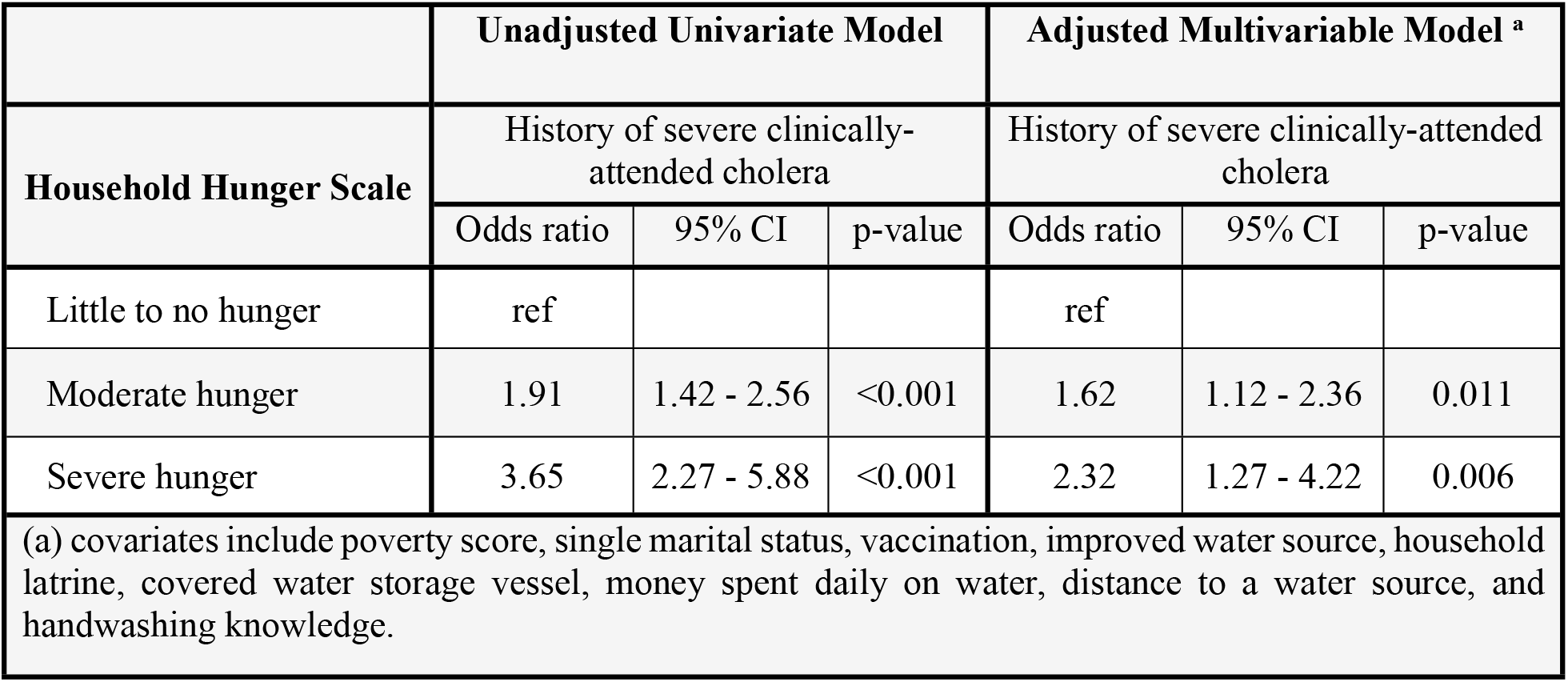
Unadjusted univariate and adjusted multivariable associations between household food insecurity and history of severe, clinically-attended cholera in a cross-sectional household survey of Mirebalais, Haiti 2019.

In a sensitivity analysis, a likelihood ratio test suggested a linear relationship between severity of food insecurity and cholera fit the data best (p=0.9997), with the odds of cholera increasing by approximately 40% for each increase in severity of household food insecurity (AOR 1.39, 95% CI 1.06-1.84; p=0.02). A second sensitivity analysis demonstrated that inclusion of missing indicator variables for variables with missing data—specifically, latrine use, and covered storage vessel—did not alter the observed associations in our models.

### Household Food Insecurity as a Predictor of High-Risk Behaviors

Household food insecurity was positively associated with three high-risk behaviors: consumption of food or beverages prepared outside of the home (moderate: AOR 1.92, 95 CI 1.45-2.5, p<0.0001; severe: AOR 4.32, 95 CI 2.61-7.28, p<0.0001), antacid use (severe: AOR 2.51, 95 CI 1.52 - 4.19; p=0.0004), and consumption of leftover non-reheated food (severe: AOR 4.35, 95 CI 2.57 - 7.38, p<0.0001) (Table 4). Severe hunger was also positively associated with reported consumption of undercooked fish—however confidence intervals were wide.

**Table 4.** Adjusted associations between household food insecurity and behaviors in a cross-sectional study of 1072 households in Mirebalais, Haiti, 2019.

|  | Little to no Hunger |  |  | Moderate Hunger |  |  | Severe Hunger |  |  |
| --- | --- | --- | --- | --- | --- | --- | --- | --- | --- |
| **High-Risk Behavior<sup>a\*</sup>** | AOR | 95% CI | p-value | AOR | 95% CI | p-value | AOR | 95% CI | p-value |
| Antacid use | ref |  |  | 0.95 | 0.72 - 1.25 | 0.7041 | 2.51 | 1.52 - 4.19 | 0.0004 |
| Raw fruits/vegetables | ref |  |  | 0.69 | 0.52 - 0.92 | 0.0107 | 0.72 | 0.43 - 1.17 | 0.1868 |
| Undercooked fish | ref |  |  | 1.13 | 0.51 - 2.64 | 0.7610 | 2.48 | 0.71 - 7.76 | 0.1290 |
| Unable to reheat food | ref |  |  | 1.10 | 0.79 - 1.56 | 0.5569 | 4.35 | 2.57 - 7.38 | <0.0001 |
| Consumes outside foods | ref |  |  | 1.92 | 1.45 - 2.57 | <0.0001 | 4.32 | 2.61 - 7.28 | <0.0001 |
| (a) Covariates: poverty score, single marital status, cholera vaccination, and knowledge of handwashing. |  |  |  |  |  |  |  |  |  |
| * Consumed over the prior week |  |  |  |  |  |  |  |  |  |

### Association of Household Behaviors and Cholera

Figure 2 reports the associations between high-risk household behaviors and history of severe, medically-attended cholera. In adjusted analyses, we found an independent association between household history of cholera and consumption of outside food (AOR 1.68; 95% CI 1.09 – 2.61 p = 0.02) and antacid use (AOR 2.00; 95% CI 1.37 – 2.94, p = 0.0004) over the prior week. Consumption of undercooked fish was marginally associated with household history of severe medically-attended cholera.

**Fig. 2.**
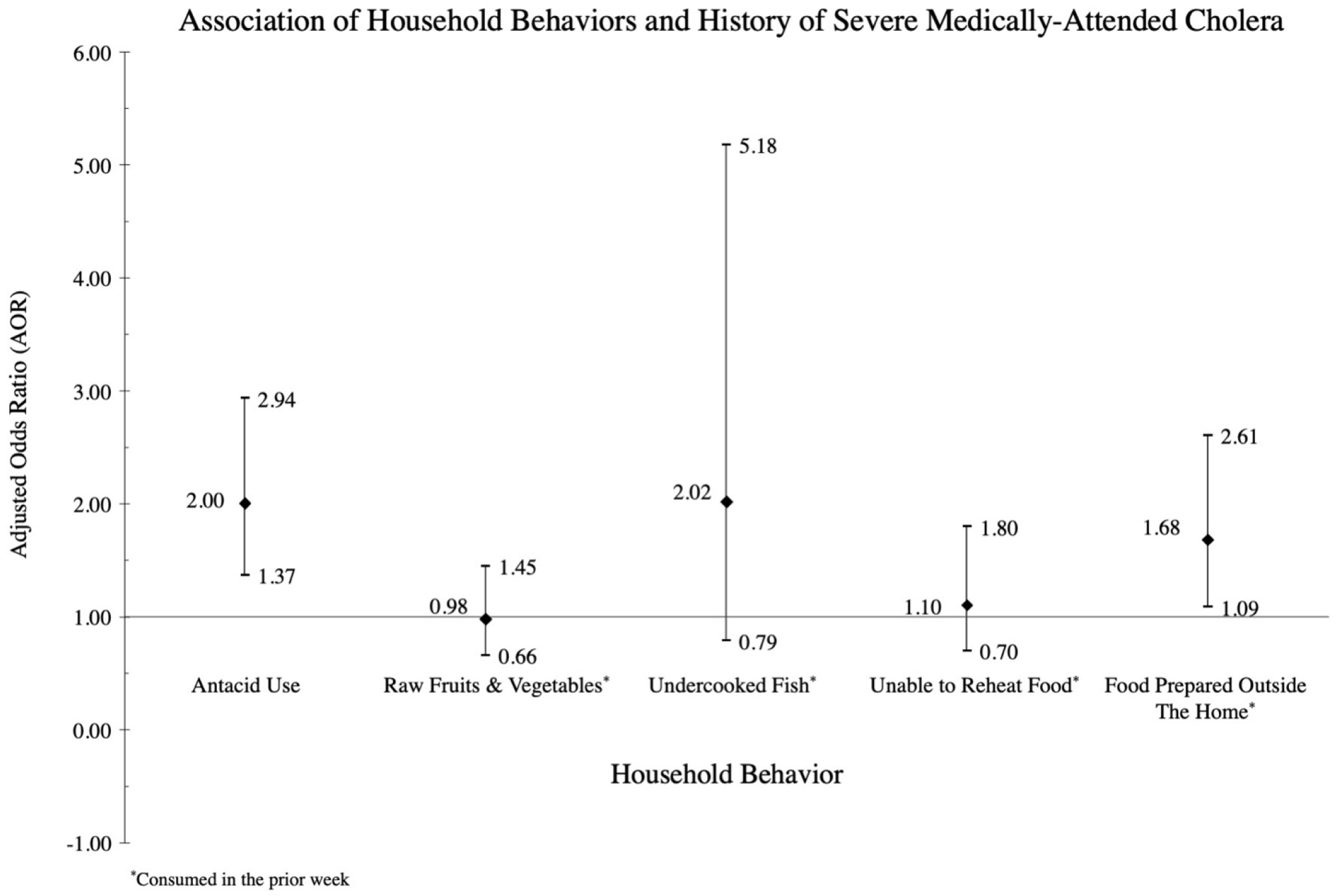
Association of high-risk behaviors and history of severe medically-attended cholera in a cross-sectional study in Mirebalais, Haiti 2019.

## Discussion

In this cross-sectional study of 1072 households in central Haiti conducted amidst a cholera outbreak, we found a positive association between food insecurity and severe medically-attended cholera and provide evidence that certain behaviors are potential mediators of this association. This study demonstrates a possible causal pathway through which food insecurity may lead to cholera.[1,2] By illuminating one pathway through which food insecurity alters cholera risk, our findings have important implications for cholera response strategies within both endemic and epidemic geographies.

We found that households with moderate and severe food insecurity were more likely to report high-risk behaviors for cholera. These included more frequent antacid use, consumption of food prepared outside the home, and eating of unheated leftover food items. Two of these behaviors—antacid use and consumption of food prepared outside the home—were also independent risk factors for cholera. A third risk factor, consumption of undercooked fish, was marginally associated with both food insecurity and cholera. These findings are consistent with a prior systematic review and metanalysis which identified all three behaviors as risk factors for cholera.[24] These behaviors likely increase the risk of infection with cholera through more frequent household contact with contaminated foods and beverages.

In both high- and low-income contexts, hunger has been shown to lead to high-risk food handling practices. One study found that individuals experiencing food insecurity in the U.S. (New Jersey) needed to make pressured choices while scavenging for meals which resulted in the consumption of contaminated food.[33] For example, participants reported consuming food left over by others as well as food items that required removal of spoiled areas with slime, mold, or insects.[33] Coping strategies are also context-specific, with insecure access to food leading to alterations in the quantity and types of foods consumed, nonfood spending allocations, and overall activity patterns.[34] Namely, prior research into informal coping strategies identified specific examples that include: “stretching and substitution techniques (such as using water in place of milk in breakfast cereals), consumption of expired and nearly expired foods, reduced meal size, meal diversity, and meal frequency, shifts to less expensive foods, and making one large pot of food to consume for several day.”

The use of antacids is a well-established risk factor for cholera and a host of other intestinal pathogens as reduced gastric acidity impairs gut immunity in hosts and has been linked with more severe symptomatic infections with cholera.[35,36] Individuals who are hungry may also experience chronic gastrointestinal complaints associated with food insecurity—as has been reported in other contexts [37–40]—that may also be relieved by antacids. Their ready availability, low cost, and utility in treating a range of symptoms make antacids a plausible first choice in response to gastrointestinal complaints among food insecure individuals, though symptomatic relief is achieved at the expense of leaving individuals more susceptible to further intestinal infections. High rates of dyspepsia and of *H. pylori* infection have been reported in Haiti and antibiotics, H_2_ blockers, and antacids are readily dispensed medications for treatment.[41,42]

The primary limitation of this exploratory study stems from its cross-sectional design, which precludes discernment of temporality. For instance, it is plausible that a household’s experience of cholera may subsequently increase their risk of food insecurity through costs associated with hospitalization, loss of income, and funerals.[18,43] While we cannot exclude the possibility that food insecurity occurred as a consequence of cholera infection, food insecurity in Haiti has existed before the cholera epidemic, and historically has been driven by high food and fuel prices, droughts, and socio-political instability.[5] Moreover, it is unlikely that cholera would directly lead to high risk household food behaviors, independently of household hunger. Regardless, the potential mediating role of food behaviors in increasing cholera risk among individuals with household hunger should be confirmed in longitudinal data.

## Conclusion

As efforts are underway to eliminate cholera as a public health problem by 2030, [44] innovative public health approaches are needed in endemic and epidemic cholera to complement the backbone of water, sanitation, hygiene and vaccination programs. This analysis furthers our understanding of the relationship between food insecurity and cholera risk. Further study should be directed toward longitudinal and qualitative studies to further explore these associations, and to elucidate whether interventions targeting food insecurity can be a complementary intervention to reduce cholera risk among populations for whom overlapping epidemics occur.

## Data Availability

The data underlying the results presented in the study are available from Louise C. Ivers.

## Acknowledgments

The authors express their sincere thanks to the study participants, the Ministry of Health and Population of Haiti, and the staff of Zanmi Lasante for their contributions to this work.

## Competing interests and financial disclosures statement

### Patient and public involvement

Our research team has conducted extensive focus group discussions and qualitative engagement on food security as a social determinant of poor health in Haiti—both published and unpublished as well as prior to and after the onset of the cholera epidemic.[27–31] These community consultations contributed to informing the premise of the analysis. Specifically, as it relates to this study, we undertook a series of community focus groups and leadership consultations related to oral cholera vaccine that contributed to our understanding of the potential connection between food security and cholera.[31] The subjects included in our study were not ‘patients’ but community members (randomly selected from our census data). Although community members themselves were not involved in the design and conduct of the study, study workers were members of the community who were trained to recruit households and administer the survey. While we have presented results of our prior cholera-related studies in community meetings, [27,30] this study which focuses on analyzing causal pathways has not been presented to community members. After publication, we will translate the abstract into Haitian Creole for more accessible dissemination.

